# Genetically Guided Precision Medicine Clinical Decision Support Tools: A Systematic Review

**DOI:** 10.1101/2023.08.23.23294506

**Authors:** Darren Johnson, Guilherme Del Fiol, Kensaku Kawamoto, Katrina Romagnoli, Nathan Sanders, Grace Isaacson, Elden Jenkins, Marc Williams

**Author notes:** Darren Johnson, 5090 South 1130 West, Taylorsville UT 84123, 8015202539.

## Abstract

**Objective:** Patient care using genetics presents complex challenges. Clinical decision support (CDS) tools are a potential solution because they provide patient-specific risk assessments and/or recommendations at the point of care. This systematic review evaluated literature on CDS systems which have been implemented to support genetically guided precision medicine (GPM).

**Materials and Methods:** A comprehensive search was conducted in MEDLINE and Embase, encompassing Jan 1^st^, 2011 to March 14^th^, 2023. The review included primary English peer-reviewed research articles studying humans, focused on use of computers to guide clinical decision making and delivering genetically guided, patient-specific assessments and/or recommendations to healthcare providers and/or patients.

**Results:** The search yielded 3,832 unique articles. After screening, 41 articles were identified that met the inclusion criteria. Alerts and reminders were the most common form of CDS used. 27 systems were integrated with the electronic health record; 2 of those used standards-based approaches for genomic data transfer. Three studies used a framework to analyze the implementation strategy.

**Discussion:** Findings include limited use of standards-based approaches for genomic data transfer, system evaluations that do not employ formal frameworks, and inconsistencies in the methodologies used to assess genetic CDS systems and their impact on patient outcomes.

**Conclusion:** We recommend that future research on CDS system implementation for genetically guided precision medicine should focus on implementing more CDS systems, utilization of standards-based approaches, user-centered design, exploration of alternative forms of CDS interventions, and use of formal frameworks to systematically evaluate genetic CDS systems and their effects on patient care.

## Background

Genetic disorders, while individually rare, are collectively common [1]. These disorders affect between 2-10% of the population, occur in all medical specialties, and are more common in specific populations [2–5]. Genetically guided precision medicine (GPM) entails the delivery of individually tailored medical care that leverages information about each person’s unique characteristics, including clinical data, genetic test results, patient preferences, and family health history (FHx) [6–8]. GPM is expanding with advances in genomic tools and decreasing genetic testing costs [9–11]. Genomic testing can facilitate diagnosis and inform condition-specific clinical care [12–15]. However, evidence indicates that providers have limited proficiency, self-efficacy, and resources to guide decisions about when and how to order genetic tests, refer to specialists, or change treatment or surveillance based on genomic information [16–18]. As a result, genomic testing in clinical care is underutilized [19–21]. The challenges facing clinicians will continue to grow as the volume of information generated expands the evidence base for linking genetic variation with human disease [22]. For GPM to achieve the potential to tailor medical treatments and therapies to the individual characteristics of each patient, new approaches are needed to support integration of genomic medicine in clinical decision making, especially given the limited genomic specialist workforce [23].

The essential role of health information technology in overcoming the barriers that GPM faces is recognized [24–26]. A key challenge is the integration of genomic data into electronic health records (EHRs), as genomics data possess unique characteristics that set them apart from other types of clinical data. Genomic data are highly complex, voluminous, and dynamic, yet germline genetic test results do not change over an individual’s lifetime. These attributes require specialized approaches for storage, retrieval, and ongoing interpretation [27, 28]. Currently, genomic data are often poorly integrated into EHRs, resulting in suboptimal clinical use.

Clinical decision support (CDS) systems have been proposed to address these barriers by facilitating the integration and utilization of data, including genomic data in clinical care [29]. CDS systems may support clinicians in ordering genetic testing and the management of care, preventing harmful or unnecessary interventions and decreasing delays in diagnosis, which can lead to excessive healthcare utilization and potentially inappropriate testing and treatment [30]. CDS systems can present information to users in a variety of ways, such as standalone systems (either electronic or paper) or systems which are integrated into the clinical workflow via the EHR [31]. CDS systems may be used asynchronously, meaning use would not be associated with a specific patient encounter, instead identifying individuals in need of a given service (e.g., vaccination, mammogram) as part of population health initiatives. CDS systems can also be used synchronously when presenting patient-specific information within the context of an encounter. By guiding assessments or recommendations at the point of care based on clinical management guidelines, best practices, and evidence [32], CDS systems have the potential to effectively influence changes in clinical care, thus facilitating the use of genomic data in clinical decision making [33].

In 2011, Welch and Kawamoto conducted a systematic review about genetic CDS systems, including prototypes and EHR-integrated CDS systems [35]. This review showed an abundance of CDS systems in prototype stage and stand alone CDS systems, with very few implemented outside of a pilot setting. Since this review, the use and evaluation of genetic CDS systems in clinical settings have proliferated, yet there has not been a review of studies summarizing the implementation, use, and evaluation of these in health care settings. To understand the evolution of the development and implementation of genomic CDS systems, we conducted a systematic review on genomic CDS systems which have been implemented in a clinical setting.

## Methods

A systematic review was conducted to integrate quantitative and qualitative evidence using the Rapid and Rigorous Qualitative Data Analysis (RADaR) technique [34] to examine elements surrounding implemented genomic clinical decision support (gCDS) tools. The study protocol was registered in PROSPERO (ID: 416709).

### Literature Search Strategy

A bioinformaticist (D.J.) consulted with a PhD researcher/medical librarian (K.M.R.) to search MEDLINE and Embase from January 1^st^, 2011 to March 14^th^, 2023 using a search strategy adapted from previous systematic reviews of CDS [35], genetic testing, genetic health services, and family history (FHx) (Supplementary appendix 1). The initial literature search was conducted on November 15, 2021 and an additional literature search was conducted on March 14, 2023 to capture additional citations. Both subject headings and text terms were used to search for CDS tools and genetic testing, including FHx. Reference lists of included articles and relevant systematic reviews were hand-searched for additional references.

### Study Selection

References were reviewed against the following inclusion criteria: primary English peer-reviewed research article studying humans, focused on the use of computers to guide clinical decision making and delivering genetically guided, patient-specific assessments and/or recommendations to healthcare providers and/or patients. Each stage of literature screening was conducted by two independent reviewers (D.J. and M.S.W./N.S./E.J./E.I./C.J./J.C.) using DistillerSR and a screening form. Titles and abstracts were screened to assess whether the articles met inclusion criteria. Conflicting decisions were discussed and resolved or moved to full text review. Full text review of included or conflicted abstracts was conducted such that articles included in the final systematic review met all inclusion criteria. Conflicted articles were discussed until consensus was reached or resolved by a third reviewer (M.S.W.).

### Data abstraction

Quantitative and qualitative data extraction were completed on each of the articles that met the inclusion criteria. Data extraction items for all articles are described in Supplementary appendix 3.

### Data Analysis

Using the abstracted attributes, the manuscripts were grouped into categories according to CDS type and clinical application area. The findings from these manuscripts were summarized through tables and narrative discussion. Notable themes and trends were identified and discussed. Reported barriers and facilitators were noted. Each article was classified according to the CDS taxonomy [36] and CDS critical success features [37]. A quantitative analysis of CDS trials to identify features critical to successful CDS tools was considered. However, due to a limited sample size of CDS trials and the lack of implementation data, this analysis was not feasible.

## Results

The initial MEDLINE and Embase searches identified 3,832 unique potentially relevant articles. The title and abstract review excluded 3,438 articles. The remaining 394 underwent full text review, in which 353 were rejected. The PRISMA diagram (Fig. 1) articulates the inclusion process. This left 41 primary research articles for analysis that were published from January 1^st^, 2011 to March 14, 2023 (Table 1) [38–78].

**Fig 1:**
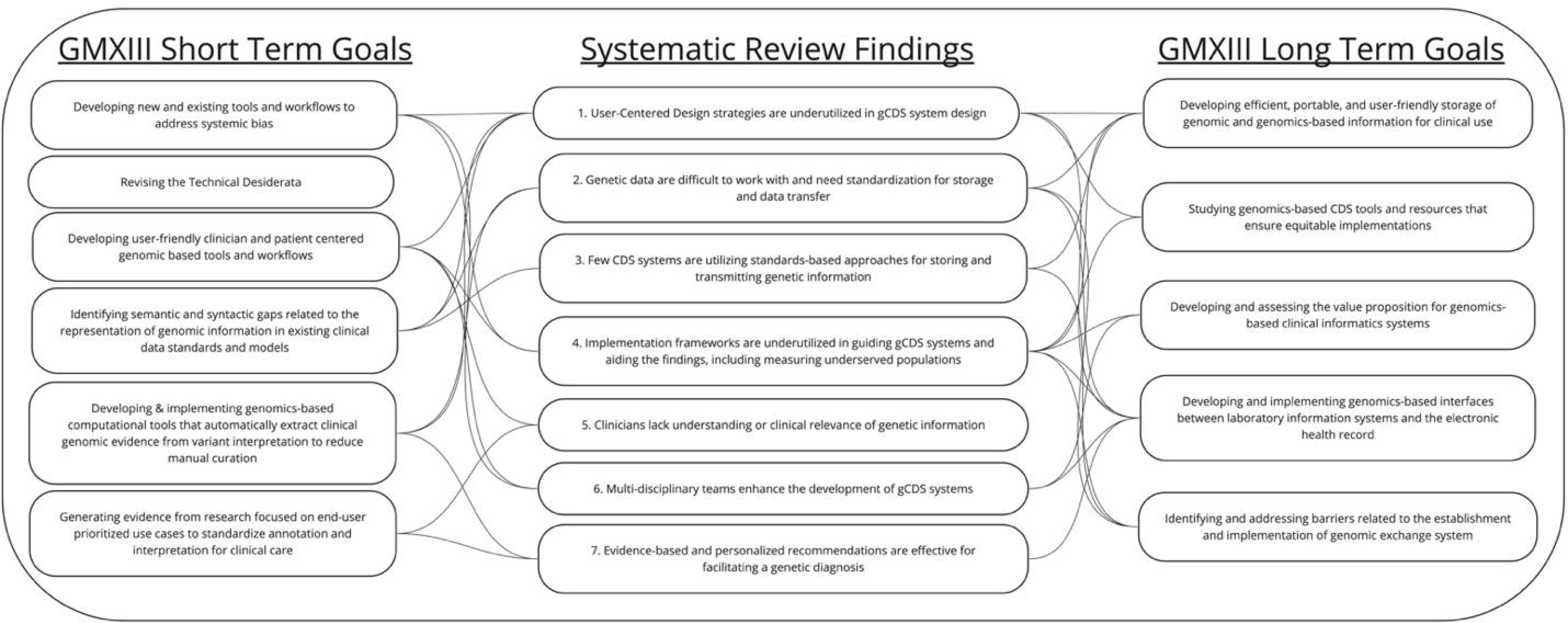
PRISMA diagram

**Table 1.**
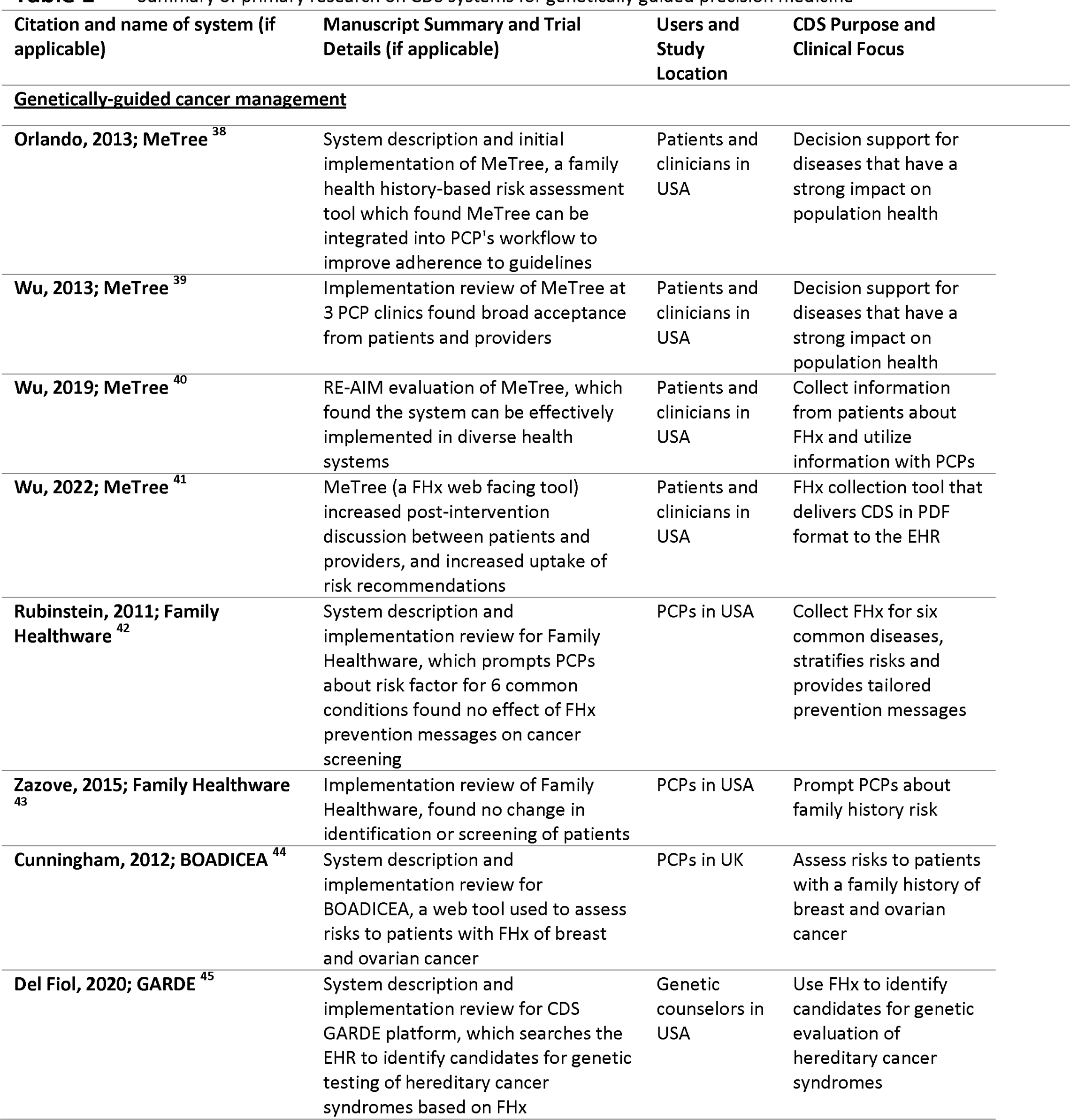
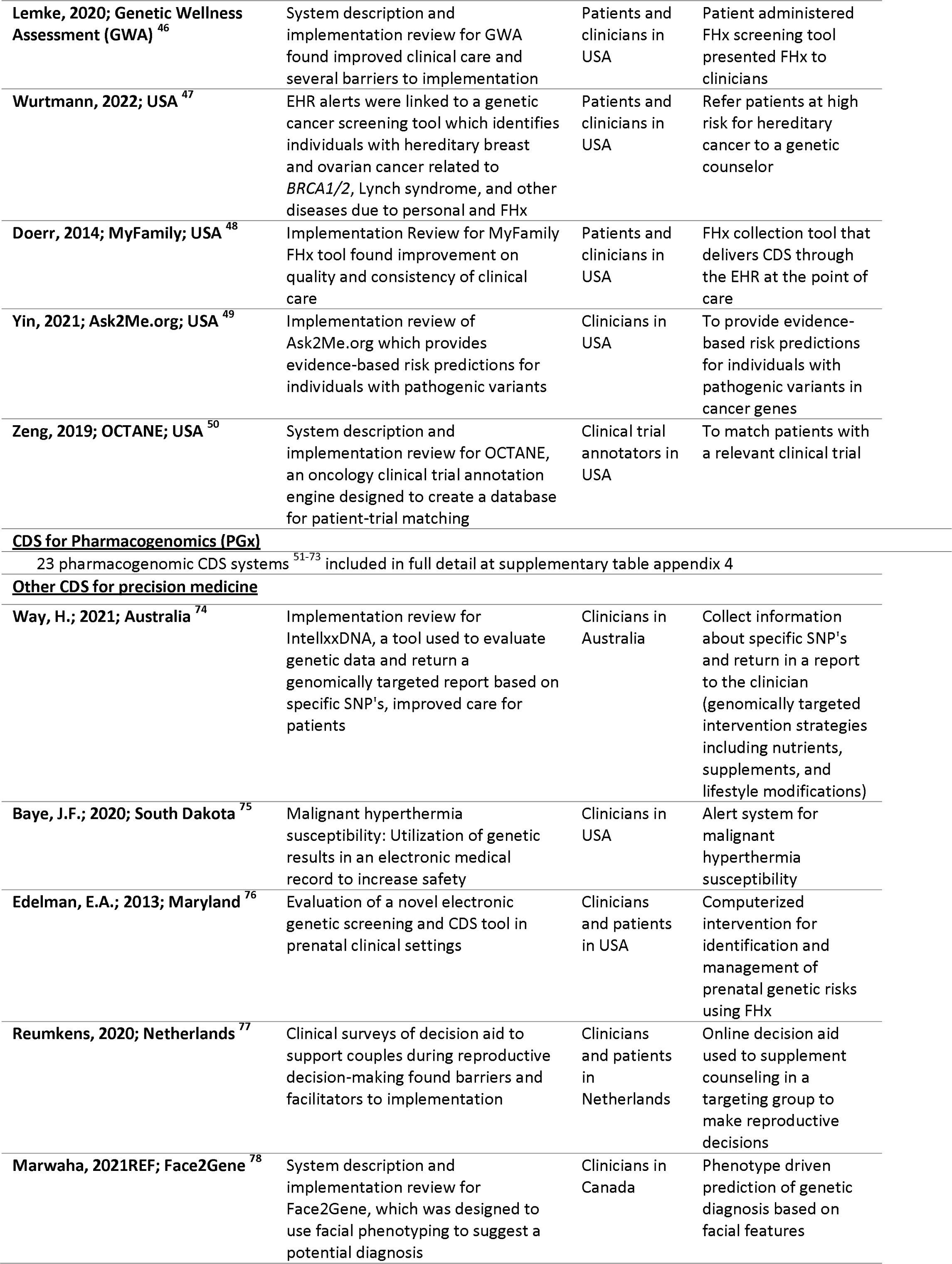
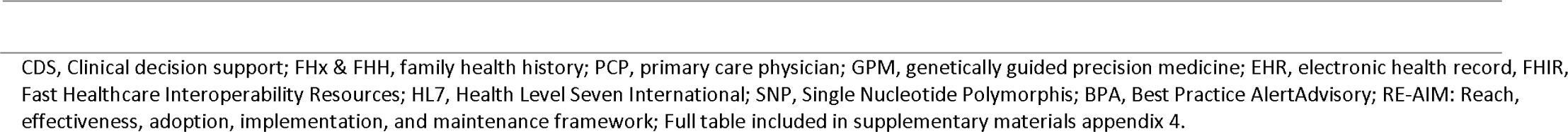
Summary of primary research on CDS systems for genetically guided precision medicine.

The described CDS tools were first categorized according to three areas of clinical focus: genetically-guided cancer management, pharmacogenomics (PGx), and other systems for precision medicine. CDS tools were further categorized by type of software architecture approach.

- Stand Alone CDS: a stand alone CDS tool which utilizes information outside of the EHR and does not interact or exchange data with the EHR (14 articles).
- EHR Proprietary: A CDS tool that works within the native EHR but does not report using industry interoperability standards for CDS or genomic data storage and transfer (25 articles).
- EHR standards-based : A CDS tool that utilizes the native EHR and transfers genetic data using standard genomic data storage and transfer methods (2 articles).

## CDS systems for genetically guided cancer management

Of the 41 articles summarized in Table 1, genetically guided cancer management was the focus of 13 articles. These included four manuscripts related to the MeTree system [38–41], a family health history FHx-based risk assessment which provides FHx-driven CDS for several conditions including cancer [42], six manuscripts on other FHx-driven CDS tools for cancer management [43–48], and single manuscripts on the following subjects: population health EHR FHx-driven CDS [49] and genetically matching clinical trial patients [50].

## CDS for pharmacogenomics

CDS to support PGx implementation was the focus of 23 of the implemented CDS systems (Table 1). Large research groups (eMERGE, PREDICT, U-PGx) composed of several institutions were responsible for 3 studies [60, 62, 66]. These papers highlighted cross institutional problems including data standards and privacy. Single site/organization implementation was described in 14 articles [50–59, 61, 64–65, 68–72] and clinical workflow and information presentation were highlighted as both barriers and facilitators. The remaining papers included implementations with a focus on the role of pharmacists [63], experiences learned from transitioning from single-gene testing to panel based testing [73], and experiences of providers with the implemented systems [67].

## Other CDS systems for genetically guided precision medicine

Five primary research articles focused on other clinical domains including two condition-specific tools designed within an institution: one directed towards anesthesiologists to help with management of patients at risk for malignant hyperthermia [75] and another for autism diagnosis and treatment [74]. A third study described a pregnancy genomic family planning tool using FHx as a driver for the CDS [76]. A fourth study using a pregnancy support tool to manage prenatal genetic risks using FHx [77]. Last, a study by Marwaha et al. described a tool using facial phenotyping to assist with diagnosis of genetic syndromes [78].

## CDS type and critical features

Point of care alerts were the most common mechanism to deliver interventions, used in 32 of the gCDS systems (Table 2). Barriers to the implementation of alerts were described in 17 of the gCDS systems including alert fatigue, difficult integration with EHR, and lack of understanding of genetic information presented with the alert. Facilitators to the implementation of alerts were also mentioned in 14 of the gCDS systems including lack of interruption of clinical workflow, integration with EHR, and access to point-of-care information. There was little work reported on designing tools to work as expert systems (e.g., diagnostic decision support) or for workflow support (e.g., support in genetic test ordering, clinical documentation). Lack of genetic data standards was mentioned several times as a barrier to multi-departmental and multi-institutional cooperation.

**Table 02.**
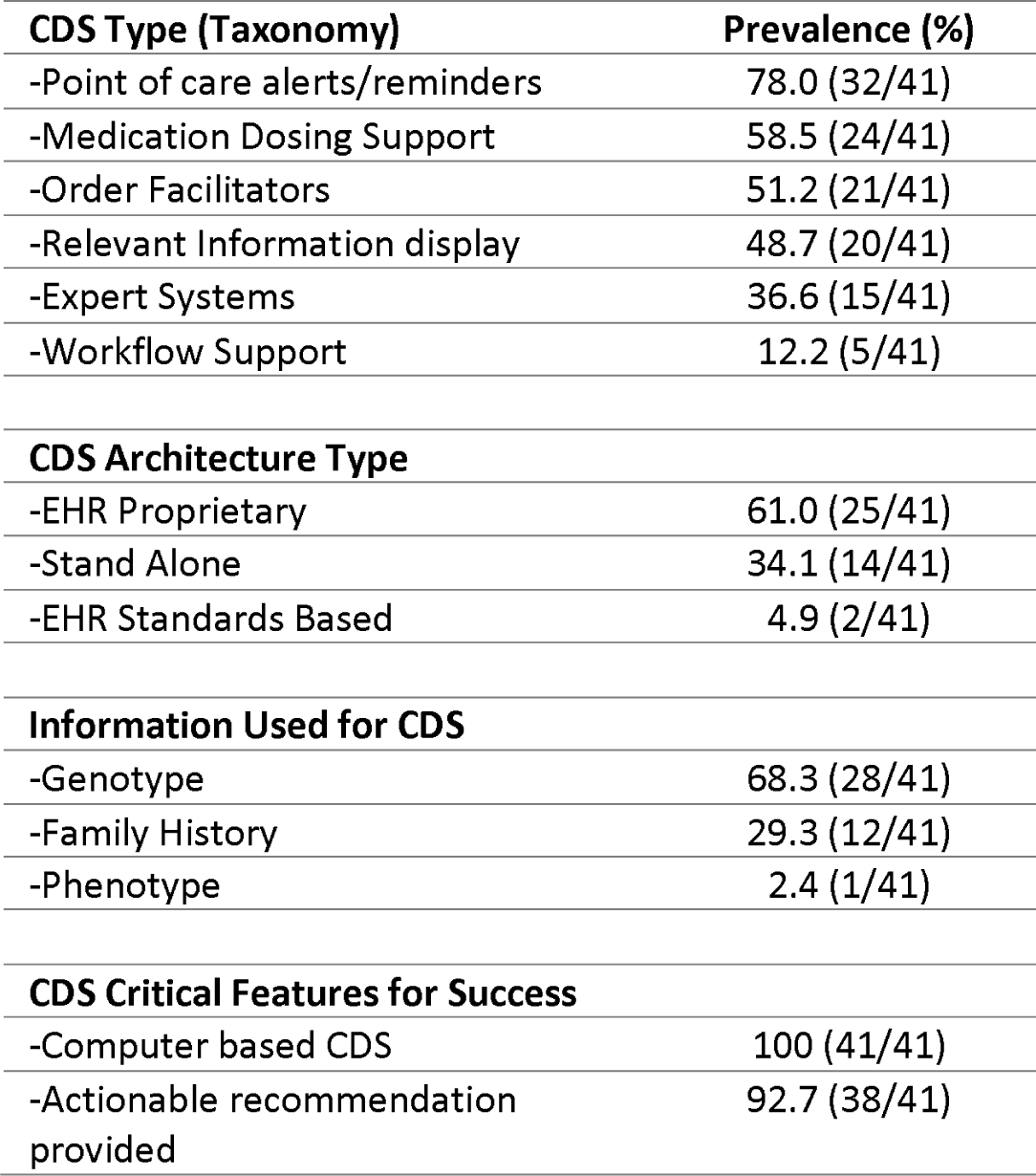
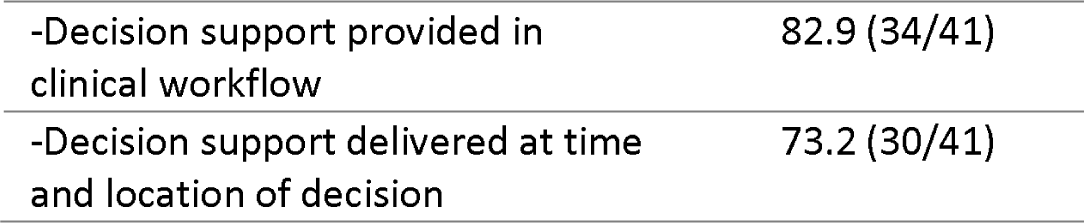
Summary of CDS taxonomy [36], CDS architecture, information used to drive CDS, and CDS critical features [37].

| CDS Type (Taxonomy) | Prevalence (%) |
| --- | --- |
| -Point of care alerts/reminders | 78.0 (32/41) |
| -Medication Dosing Support | 58.5 (24/41) |
| -Order Facilitators | 51.2 (21/41) |
| -Relevant Information display | 48.7 (20/41) |
| -Expert Systems | 36.6 (15/41) |
| -Workflow Support | 12.2 (5/41) |
| **CDS Architecture Type** |  |
| -EHR Proprietary | 61.0 (25/41) |
| -Stand Alone | 34.1 (14/41) |
| -EHR Standards Based | 4.9 (2/41) |
| **Information Used for CDS** |  |
| -Genotype | 68.3 (28/41) |
| -Family History | 29.3 (12/41) |
| -Phenotype | 2.4 (1/41) |
| **CDS Critical Features for Success** |  |
| -Computer based CDS | 100 (41/41) |
| -Actionable recommendation provided | 92.7 (38/41) |
| -Decision support provided in clinical workflow | 82.9 (34/41) |
| -Decision support delivered at time and location of decision | 73.2 (30/41) |

## Standards in genetic support systems

Of the 41 articles, only 3 reported using standards-based approaches for storing and transmitting genetic information: GARDE (FHx information using FHIR) [45], U-PGx (allowed information to be transmitted via several standards including FHIR and HL7) [60], and eMERGE (HL7 Version 2) [62]. All other systems either used proprietary approaches within vendor-based EHRs (e.g., Epic, Cerner) for data transfer, or created a unique local data repository external to the EHR as a part of the CDS architecture. 27 systems were integrated with the EHR, 3 of which reported a standards-based approach. The remaining 14 systems were considered stand-alone CDS systems.

## User centered design for genetic clinical decision support systems

Of the 30 user-centered design strategies from implementation experts [79], there were only two studies (Table 3) that adopted over 50% of the strategies (i.e., define target user, examine automatically generated data). Many studies did not report any category of user-centered design, potentially indicating a lack of understanding or use of user-centered design principles. Researchers may not be reporting on processes which help design the CDS prior to implementation.

**Table 03.** Summary of user centered design strategies for genetic CDS [79].

| User Centered Design Strategy | Percentage (%) |
| --- | --- |
| Define target users and their needs | 85.4 (35/41) |
| Examine automatically generated data | 80.4 (33/41) |
| Engage in live prototyping | 39.0 (16/41) |
| Engage in cycles of rapid prototyping | 29.3 (12/41) |
| Apply task analysis to user behavior | 24.3 (10/41) |
| Engage in iterative development | 12.2 (5/41) |
| Use generative object-based techniques | 12.2 (5/41) |
| Conduct experience sampling | 12.2 (5/41) |
| Design in teams | 9.8 (4/41) |
| Conduct heuristic evaluation | 9.8 (4/41) |
| Conduct usability tests | 9.8 (4/41) |
| Build a user-centered organizational culture | 9.8 (4/41) |
| Conduct interviews about user perspectives | 7.3 (3/41) |
| Conduct interpretation sessions with stakeholders | 4.9 (2/41) |
| Conduct focus groups about user perspectives | 4.9 (2/41) |
| Conduct design charrette sessions with stakeholders | 4.9 (2/41) |
| Collect quantitative survey data on potential users | 4.9 (2/41) |
| Define workflows | 2.4 (1/41) |
| Conduct observational field visits | 2.4 (1/41) |
| Prepare and present user research reports | 2.4 (1/41) |
| Recruit potential users | 2.4 (1/41) |
| Use associative object-based techniques | 2.4 (1/41) |
| Use dialogic object-based techniques | 2.4 (1/41) |
| Conduct artifact analysis | 0 (0/41) |
| Conduct co-creation sessions | 0 (0/41) |
| Conduct competitive user experience research | 0 (0/41) |
| Develop a user research plan | 0 (0/41) |
| Develop experience models | 0 (0/41) |
| Develop personas and scenarios | 0 (0/41) |
| Apply process maps to system-level behavior | 0 (0/41) |

## Implementation Framework

Of the 41 articles, 2 described the use of an implementation framework to develop, implement, and evaluate the CDS system (Table 4). Implementation frameworks used included RE-AIM [39–41] and PRISM [56]. MeTree utilized RE-AIM to evaluate uptake and implementation processes. Pre-implementation measures included site visits, staff surveys, and qualitative interviews to assess for readiness and identify barriers and facilitators. MeTree evaluated the population representation to measure effects on all races within the community. Surveys and interviews to assess adoption of MeTree showed positive responses to change commitment but low positive levels of change efficacy [38–41]. Implementation surveys helped identify needs for patients to understand how to access laboratory test results. Aquilante’s biobank research utilized the PRISM methodology and praised the ease of use for the final CDS system, even though the process took several years of multi-stakeholder engagements, analysis of clinical workflows, and iterative designs [56]. Other articles described implemented CDS systems with no formal application of implementation frameworks for measuring outcomes related to implementation.

**Table 04.** Summary of implementation frameworks utilized for genetic clinical decision support systems.

| Citation and name of system (if applicable) | Manuscript Summary and Trial Details (if applicable) | Implementation Framework used |
| --- | --- | --- |
| Orlando; 2013 REF; MeTree | System description and initial implementation of MeTree, a family health history-based risk assessment tool which found MeTree can be integrated into PCP's workflow to improve adherence to guidelines | RE-AIM: Reach, Adoption, Implementation, Maintenance |
| Aquilante, C.; 2020; Colorado | Implementation review for biobank PGx returning results pre-emptively to institutions | PRISM: Practical, Robust Implementation and Sustainability Model |

## Facilitators and barriers for genetic CDS implementation

The facilitators for gCDS systems include implementation of point of care alerts (29 articles), genetic data integration within EHR (23 articles), providing evidence-based and personalized recommendations for diagnosis (21 articles), multi-disciplinary teams developing the CDS (12 articles), patient-facing component of CDS (9 articles), guideline accessibility (6 articles), institutional support (5 articles), and physician champions (5 articles).

The barriers identified in the review include lack of standards for storing and transmitting genetic information (26 articles), lack of genetic data integration within the EHR (20 articles), lack of understanding of genetic information by the clinician (18 articles), alert fatigue (16 articles), cost to healthcare system with inadequate information or analysis on cost-benefit (15 articles), clinical workflow disruptions (14 articles), lack of access to genetic personnel (7 articles), and lack of patient engagement materials (4 articles).

## Trend analysis

The publication volume on CDS for GPM was relatively constant throughout the timeline of the review. The major foci of the literature in this domain have been on pharmacogenomics, FHx CDS, and CDS for cancer management.

## Discussion

As the field of precision medicine diagnostics continues to grow rapidly, there is a pressing need for more research on how to effectively harness CDS to integrate these discoveries into everyday clinical practice. For instance, even in the realm of FHx-driven CDS, which is arguably the most established area of research concerning CDS for GPM, implementations of FHx-driven CDS tools have primarily focused on hereditary cancer management. Genotype driven tools have primarily focused on PGx applications. This indicates that there is still a considerable knowledge gap in applying genotype and FHx data for GPM in many clinical specialties. Within this review, there were only 5 implementations not related to cancer or PGx. The topics of these implementations are not trivial, relating to pregnancy and family planning, autism, using phenotype information to predict genetic disease, and malignant hyperthermia risk. Given the limited literature available on any specific topic within this field, it is crucial to invest in further research to expand and deepen our understanding of what generalizable principles for gCDS can be utilized to facilitate implementation across a broad set of GPM domains.

Clinicians without formal training in genetics are usually the first to encounter patients with genetic conditions. gCDS can help clinicians recognize a genetic condition and initiate condition-specific management. gCDS systems can potentially improve and support uptake of genetic services, but the impact on influencing clinical decisions has not been measured well. The results of our systematic review indicate that while there is growing interest in implementing gCDS systems for GPM, major challenges still need to be addressed including the lack of use of standards-based approaches to integrate CDS with EHR systems, limited clinical trials using a rigorous study design, and underuse of implementation frameworks and outcomes assessment in the evaluation of CDS implementations.

We observed a strong reliance on point of care alerts as the primary CDS type. Studies reported alerts as both facilitators and barriers to successful implementation. The characteristics that facilitate implementation include proper integration into the workflow (MeTree [38–41], GatorPGx [55], Genetic Wellness Assessment [46]) and minimization of interruptions (MeTree). Interruptive alerts are often viewed as a barrier, whereas those that seamlessly integrate into the workflow act as facilitators. The lack of more sophisticated CDS, such as expert systems and CDS that provide workflow support, speaks to the difficulty in creating an effective multi-disciplinary team which would have the knowledge necessary to design the systems. However, the high rate of usage for successful gCDS features would indicate the previous research done by Kawamoto et al. [37] on the topic may have been effective in reaching those designing CDS tools.

The lack of user-centered design reported in the articles is noteworthy given the necessity of user input and feedback throughout the iterative process of creating a successful gCDS system. Our review found that few gCDS tools evaluate workflow integration through clinician input to the process, or conducted usability tests. Future studies should follow user-centered design approaches in the design and implementation of gCDS. This recommendation was reached independently by participants in the Genomic Medicine XIII meeting (GMXIII) that proposed a research agenda to support the development and implementation of genomics-based clinical informatics tools and resources [80]. One of the meeting’s short term research priorities was “Developing user-friendly clinician- and patient-centered genomics-based tools and workflows”. By emphasizing strategies that have a more positive impact, such as integrating alerts and reminders into the workflow and utilizing user-centered design principles, the effectiveness of CDS tools can be enhanced ultimately improving patient outcomes in GPM. User-centered design can also aid integration, allowing gCDS tools to be compatible with existing health information systems, such as EHRs, laboratory information systems, and fit within the clinical workflow. User-centered design principles can help to establish the specific needs for data exchange and system interfaces, enabling seamless integration and promoting the effective use of genomic data across the healthcare ecosystem. In addition, these design principles promote evidence-based practice by encouraging the development of gCDS systems based on the best available evidence.

Implementation frameworks can also play a role in the development of CDS systems for GPM. These frameworks provide a structured approach that guides the design, integration, and evaluation of gCDS systems within the healthcare setting. Studies that used implementation framework [38–41, 56] facilitated stakeholder engagement by identifying and engaging key stakeholders, including healthcare providers, patients, administrators, and IT professionals, in the development and adoption of gCDS tools. This collaborative approach ensures that the system meets the needs of all users and promotes its acceptance and utilization. gCDS systems which didn’t use implementation frameworks may have resolved specific barriers mentioned in the gCDS systems with their use, but this could not be assessed from the published results.

Implementation frameworks also facilitate evaluation and continuous improvement by providing a roadmap for assessing the impact of gCDS tools on clinical practice, patient outcomes, and healthcare costs. This evaluation process enables ongoing refinement and improvement of the system, ensuring that it continues to meet the evolving needs of clinicians and patients. The need for evaluation of the implementation of gCDS and other information systems to support GPM was endorsed in both the short and long-term research recommended by GMXIII. [80]

Our review revealed that the lack of use of standards for genomic data integration and representation remains a barrier to the incorporation of genetic information into EHRs and gCDS systems. Despite the previously highlighted importance of adopting standards [27, 28] including prioritization in the recommendations from GMXIII, they are not being widely incorporated. Data standardization is mentioned in nearly all of the implementations, yet only three mentioned a specific data standard. Several articles mentioned the cost and resource constraints associated with implementing standards could deter smaller institutions from adopting them [49,53,56,60]. The issue of resource constraints affecting equitable implementation of gCDS and other information systems was highlighted in the recommendations of GMXIII. Implementations like the GatorPGx [55] and Family Healthware [42, 43] call for standards as a way to promote data accuracy and consistency. Multi-institution projects (U-PGx, PREDICT, eMERGE) also cited data standards as a way to enable the scalability of gCDS systems, allowing them to be more easily implemented in diverse healthcare settings.

## Strengths and limitations

Use of a validated and systematic approach to the literature review was the major strength of this study. While the conclusions are necessarily subjective, they correspond to recommendations for the research agenda independently developed through an expert consensus process at the GMXIII meeting. In addition to the mentions in the text, Figure 2 maps the conclusions from the review to the recommendations from GMXIII.

**Fig 2:**
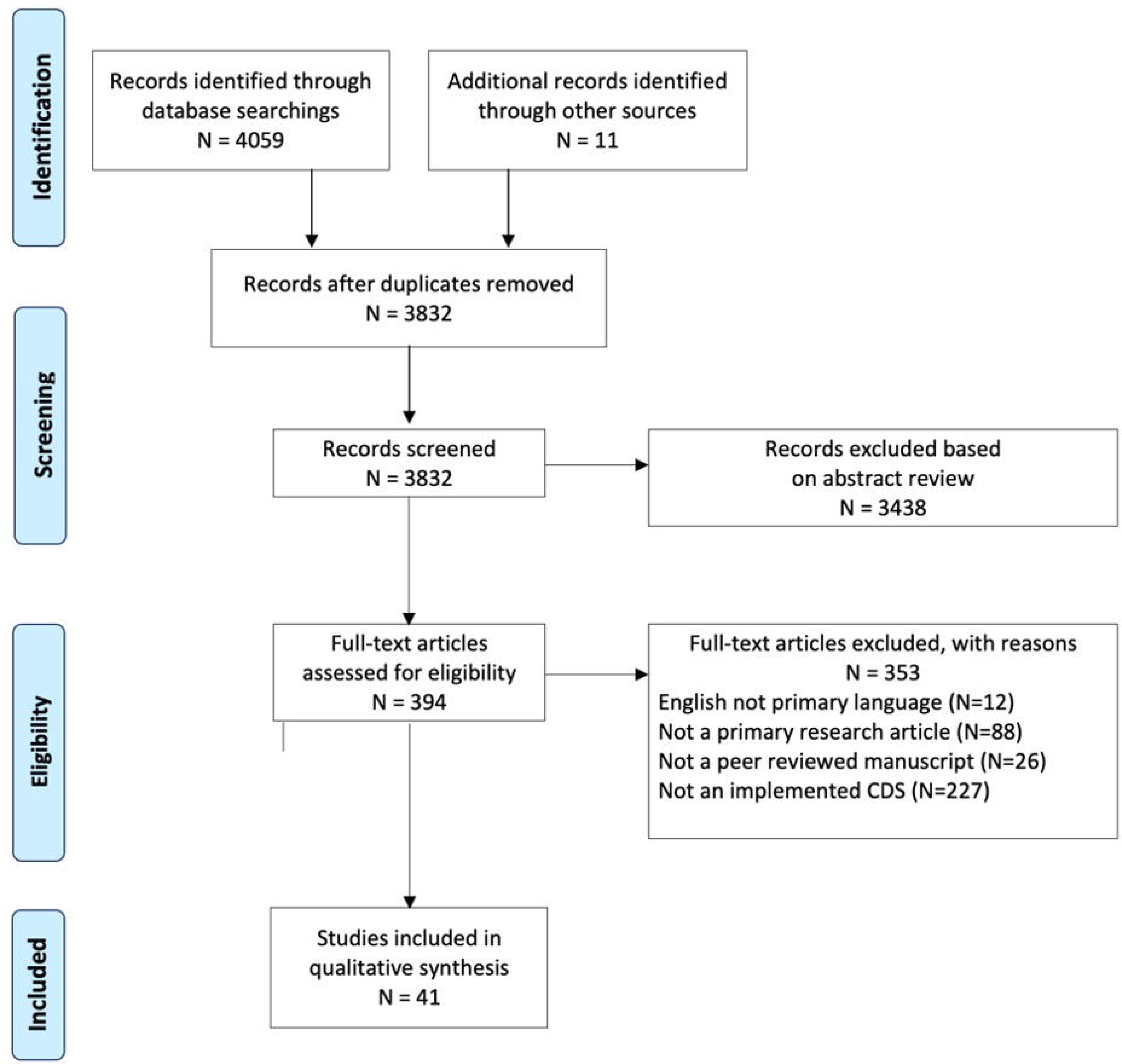
GMXIII recommendations, systematic review mapping results to recommendations.

In terms of limitations, this study does not provide a quantitative meta-analysis of the impact of CDS interventions for GPM. A meta-analysis was not possible due to the limited number of outcome studies in this field and the heterogeneous nature of the various interventions and clinical domains. Second, we only included manuscripts written in English, which may have led to some relevant manuscripts being excluded. Third, we only looked at published manuscripts, which excluded any tools that were developed for the commercial market and have been implemented but may not have published results of any evaluations that performed on these systems.

## Conclusion

Genetic disorders affect a significant portion of the population and present challenges for healthcare providers who may lack the necessary proficiency or confidence to effectively utilize genetic testing in clinical care. As genomic CDS tools advance and the cost of genetic testing decreases, GPM offers the potential to tailor medical treatments to individual patient characteristics. However, barriers persist in integrating genomic data into EHRs and CDS systems, which are critical for managing genetic testing and optimizing patient outcomes. Our systematic review of gCDS systems demonstrates that more systems are being implemented in healthcare systems, but also highlights the need for improvements in the integration of standard genomic data and the evaluation of these systems to maximize their potential in clinical practice.

To address these challenges, future research is needed to explore the use of user-centered design, unified standards-based approaches for developing gCDS tools, and implementation frameworks in the design, implementation, and evaluation of these tools. By promoting stakeholder engagement, data management, interoperability, evidence-based practice, and continuous improvement [38–41, 56], implementation frameworks can facilitate the successful adoption and utilization of genomic CDS tools in healthcare settings ultimately ensuring healthcare professionals will be better equipped to make informed decisions and improve patient care by incorporating the latest advancements in genomics into their practice.

## Data Availability

All data produced in the present study are available upon reasonable request to the author

## ACKNOWLEDGEMENTS

We thank Dr. Raghuveer Puttagunda (Geisinger Health Systems), Connor Jenkins (Rocky Vista Medical School) for their abstract review. We also thank Zachari Salvati (Geisinger Health Systems) for his continual input and review of the manuscript.

The authors have no competing interests to disclose

This project was funded by a grant from the National Institute of Health’s National Human Genome Research Institute R01HG011799

## Notes

### Competing Interest Statement

The authors have declared no competing interest.

### Funding Statement

This project was funded by a grant from the National Institute of Healths National Human Genome Research Institute R01HG011799

