## Supplementary Appendix 1 for "Genetically Guided Precision Medicine Clinical Decision Support Tools: A Systematic Review"

**Supplemntary appendix 1 (Search Strategy)**

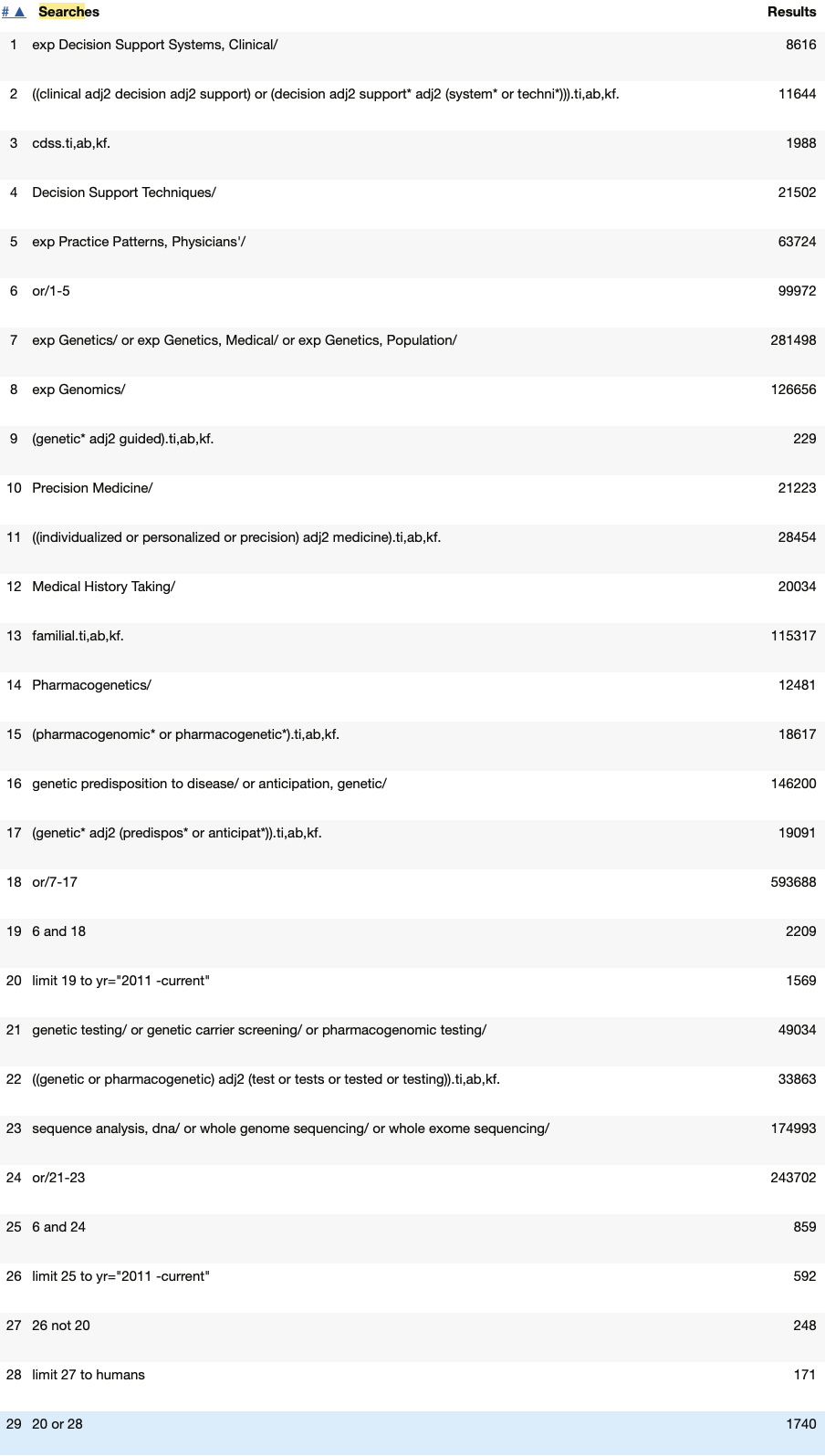

EMBASE search strategy

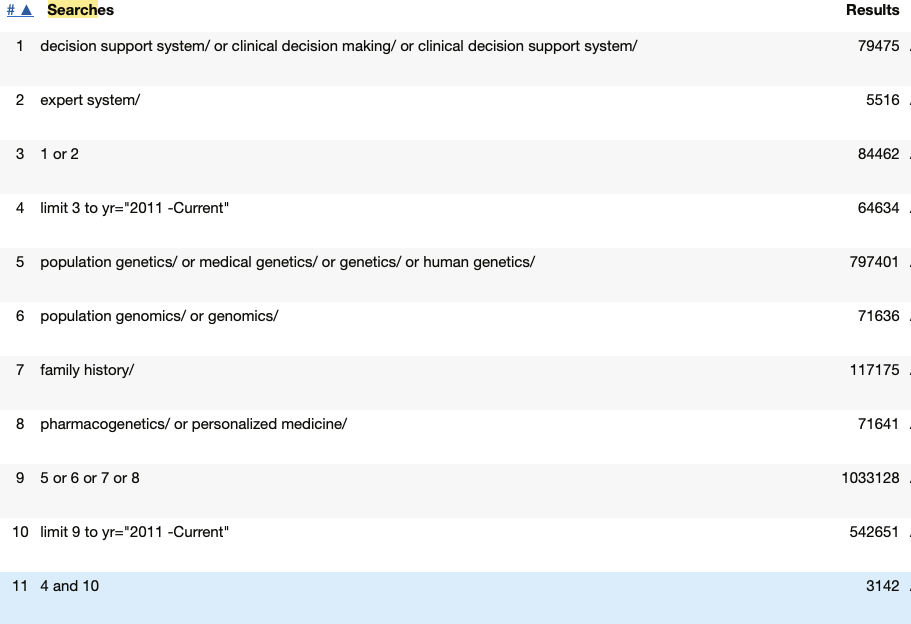

**Supplementary appendix 2 (manuscript inclusion criteria)**

Definitions

- Healthcare Provider: Physician, nurse practitioner, physician assistance, registered nurse, genetic counselor
- Genetic Factor: genotype, gene expression profile, and/or family health history
- EHR Proprietary: A CDS tool that works within the native electronic health record but does not report using industry standards surrounding genomic data storage and transfer
- EHR Standards-based: A CDS tool that utilizes the native electronic health record and transfers genetic data using standard genomic data storage and transfer methods
- Stand Alone CDS: a CDS tool which utilizes information outside of the electronic health record and does not interact or exchange data with the EHR

Additional inclusion criteria (at least one):

- Intervention study evaluating the impact of a CDS system in an actual patient care context
  - For a comparative intervention study, CDS required to be a part of the primary intervention under evaluation
  - Excludes laboratory evaluations or simulations

Methodology article whose primary focus is on how CDS systems should be designed specifically to support clinical delivery of patient0specific assessments and/or recommendations guided by genetic factors. Includes system description articles

**Supplementary appendix 3**

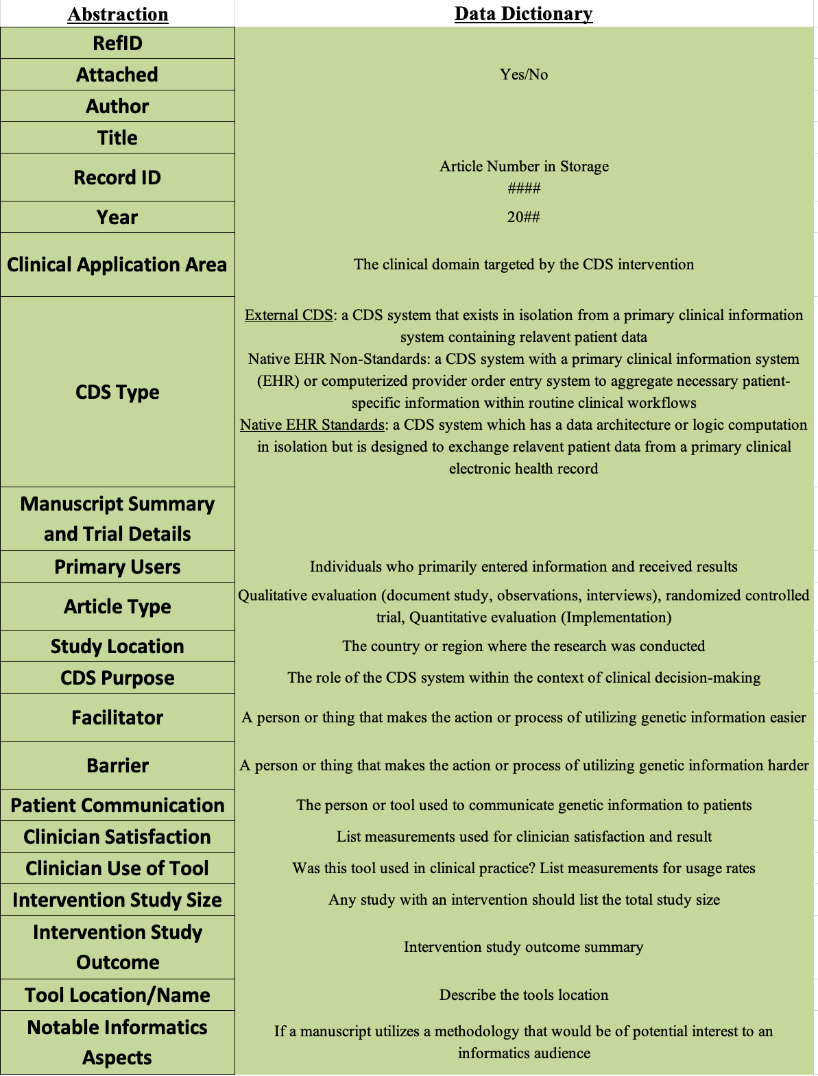

**Supplementary Appendix 4**

Full table of papers included in Systematic Review

Summary of primary research on CDS systems for genetically guided precision medicine

| Citation and name of system (if applicable) | Manuscript Summary and Trial Details (if applicable) | Users and Study Location | CDS Purpose and Clinical Focus |
| --- | --- | --- | --- |
| Genetically-guided cancer management | | | |
| Orlando, 2013; MeTree ^38^ | System description and initial implementation of MeTree, a family health history-based risk assessment tool which found MeTree can be integrated into PCP's workflow to improve adherence to guidelines | Patients and clinicians in USA | Decision support for diseases that have a strong impact on population health |
| Wu, 2013; MeTree ^39^ | Implementation review of MeTree at 3 PCP clinics found broad acceptance from patients and providers | Patients and clinicians in USA | 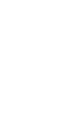Decision support for diseases that have a strong impact on population health |
| Wu, 2019; MeTree ^40^ | RE-AIM evaluation of MeTree, which found the system can be effectively implemented in diverse health systems | Patients and clinicians in USA | Collect information from patients about FHx and utilize information with PCPs |
| Wu, 2022; MeTree ^41^ | MeTree (a FHx web facing tool) increased post-intervention discussion between patients and providers, and increased uptake of risk recommendations | Patients and clinicians in USA | 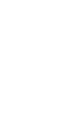FHx collection tool that delivers CDS in PDF format to the EHR |
| Rubinstein, 2011; Family Healthware ^42^ | System description and implementation review for Family Healthware, which prompts PCPs about risk factor for 6 common conditions found no effect of FHx prevention messages on cancer screening | PCPs in USA | Collect FHx for six common diseases, stratifies risks and provides tailored prevention messages |
| Zazove, 2015; Family Healthware ^43^ | Implementation review of Family Healthware, found no change in identification or screening of patients | PCPs in USA | Prompt PCPs about family history risk |
| Cunningham, 2012; BOADICEA ^44^ | System description and implementation review for BOADICEA, a web tool used to assess risks to patients with FHx of breast and ovarian cancer | PCPs in UK | Assess risks to patients with a family history of breast and ovarian cancer |
| Del Fiol, 2020; GARDE ^45^ | System description and implementation review for CDS GARDE platform, which searches the EHR to identify candidates for genetic testing of hereditary cancer syndromes based on FHx | Genetic counselors in USA | Use FHx to identify candidates for genetic evaluation of hereditary cancer syndromes |
| Lemke, 2020; Genetic Wellness Assessment (GWA) ^46^ | System description and implementation review for GWA found improved clinical care and several barriers to implementation | Patients and clinicians in USA | Patient administered FHx screening tool presented FHx to clinicians |
| Wurtmann, 2022; USA ^47^ | EHR alerts were linked to a genetic cancer screening tool which identifies individuals with hereditary breast and ovarian cancer related to *BRCA1/2*, Lynch syndrome, and other diseases due to personal and FHx | Patients and clinicians in USA | Refer patients at high risk for hereditary cancer to a genetic counselor |
| Doerr, 2014; MyFamily; USA ^48^ | Implementation Review for MyFamily FHx tool found improvement on quality and consistency of clinical care | Patients and clinicians in USA | FHx collection tool that delivers CDS through the EHR at the point of care |
| Yin, 2021; Ask2Me.org; USA ^49^ | Implementation review of Ask2Me.org which provides evidence-based risk predictions for individuals with pathogenic variants | Clinicians in USA | To provide evidence-based risk predictions for individuals with pathogenic variants in cancer genes |
| Zeng, 2019; OCTANE; USA ^50^ | System description and implementation review for OCTANE, an oncology clinical trial annotation engine designed to create a database for patient-trial matching | Clinical trial annotators in USA | To match patients with a relevant clinical trial |
| CDS for Pharmacogenomics (PGx) | | | |
| Gill, P.S.; 2021; Arkansas ^51^ | System description of EPIC's genomic indicators to return results for a clinical PGx test | Clinicians in USA | To provide an information alert to clinicians about potential PGx drug interactions |
| Manzi, S.F.; 2016; Boston ^52^ | System description and implementation review for system which prompts clinicians based on laboratory results of PGx test found improved clinical outcomes | Clinicians in USA | To provide information about PGx to clinician at time of patient interaction at time of order |
| Hinerer, M; 2017; Germany ^53^ | Clinical surveys of sites with PGx implementation reviewed barriers and facilitators for implementing PGx | Clinicians in Germany | PGx CDS evaluation at 8 German hospitals |
| Zastrozhin, M; 2020; Russia ^54^ | Implementation of www.pgx2.com as a PGx interpretation tool found improved dosing and reduced side effects | Clinicians in Russia | Interpret and display results of PGx testing |
| Marrero, R.; 2020; GatorPGx ^55^ | Implementation review of GatorPGx which provides recommendations for PGx implementation | Clinicians in USA | Preemptive PGx testing alerts clinicians at point of care |
| Aquilante, C.; 2020; Colorado ^56^ | Implementation review for biobank PGx returning results preemptively to institutions | Clinicians and patients in USA | Preemptive PGx testing alerts clinicians at point of care |
| Borden, B.; 2018; GPS; USA ^57^ | Implementation review for Genomic Prescribing System PGx results found implementation barriers and increased utilization of PGx information | Clinicians in USA | Delivering PGx alerts to clinicians |
| Hernandez, W.; 2020; GPS; USA ^58^ | Implementation review for GPS PGx found genetic ancestry specific care improvements | Clinicians in USA | Using genetic ancestry to enable pre-emptive PGx alerts |
| Petry, N.; 2019; South Dakota ^59^ | System description and implementation review of EPIC BPA's for PGx information delivery found improved care with barriers to implementation | Clinicians in USA | Preemptive PGx testing alerts clinicians at point of care |
| Blagec, K.; 2018; Austria ^60^ | Implementation review of U-PGx GIMS which manages genetic information and communicates results found improved care through several complementary methods (paper, electronic, and mobile) | Clinicians in Austria | Enable PGx CDS by secure transfer of PGx test results with dosing recommendations, generation of a PGx report, and a safety card enabling mobile based PGx CDS |
| Danahey, K.; 2017; Illinois ^61^ | System description and initial implementation of Genomic Prescribing System (GPS) found improved information delivery for PGx information to clinicians | Clinicians in USA | An online, secure, electronic custom interface for storing and delivering point-of-care genomic information |
| Rasmussen-Torvik, L.; 2015; USA ^62^ | System description and initial implementation of eMERGE-PGx project for preemptive PGx in electronic health records | Clinicians in USA | Preemptive PGx testing alerts clinicians at point of care |
| Osusu-Obeng, A.; 2014; Florida ^63^ | Roles for pharmacists emerged during clinical implementation of genotype-guided clopidogrel therapy | Clinicians in USA | Genotype drives EHR alerts with patient-specific genotype-guided drug therapy recommendations |
| Bielinski, S.J.; 2014; Minnesota ^64^ | Preemptive PGx testing for biobank participants included targeted sequencing of 84 PGx genes. Synchronous real-time CDS is integrated in the EHR, flags drug-gene interactions and provides recommendations | Clinicians in USA | Preemptive PGx testing alerts clinicians at point of care |
| Goldspiel, B.R.; 2013; Maryland ^65^ | A PGx committee was established to develop CDS algorithms for medications with hypersensitivity reactions at the point of care. Prescribers overrode recommendations mainly due to previous tolerance | Clinicians in USA | Preemptive PGx testing alerts clinicians at point of care |
| Pulley, J.M.; 2012; Tennessee ^66^ | Operational implementation of prospective genotyping linked to an advanced CDS system. This included patient and clinician engagement | Clinicians in USA | Preemptive PGx testing alerts clinicians at point of care |
| Carabello, P.; 2017; Minnesota ^67^ | A comprehensive operational model can support PGx implementation. Institutions can use this model as a roadmap to support similar efforts | Clinicians in USA | Preemptive PGx testing alerts clinicians at point of care |
| Obeng, A.O.; 2016; New York ^68^ | Development of a strategy that accounts for the complexity of race and leverages EHRs for algorithm variables and deploying point-of-care dose recommendations | Clinicians in USA | Preemptive PGx testing alerts for warfarin clinicians at point of care |
| Ramsey, L.B.; 2017; Ohio ^69^ | Implementation of PGx at Cincinnati Children’s Hospital: the development, launch, lessons learned and updates to the system after 14 years | Clinicians in USA | Preemptive PGx testing alerts clinicians at point of care |
| Bell, G.C.; 2014; Tennessee ^70^ | Active CDS delivered through an EHR facilitates gene-based prescribing and other application of genomics to patient care. Interruptive CDS appropriately guided prescribing in 95% of patients for whom they were issued | Clinicians in USA | Preemptive PGx testing alerts clinicians at point of care |
| Wick J.A.; 2014; Ohio ^71^ | Implementation of PGx across 30 primary care sites led to changes in recommended drugs 82% of the time, preventing adverse drug events and increasing quality of care | Clinicians in USA | CDS indicates which patients meet criteria for PGx testing |
| Peterson, J.F.; 2013; Tennessee ^72^ | The design of a locally developed EHR supporting PGx has a generalizable utility. Genetic data from panel-based genotyping were sequestered from the EHR until drug-gene interactions met evidentiary standards. | Clinicians in USA | Preemptive and indication triggered PGx testing alerts clinicians at point of care |
| Cicali, E.J.; 2019; Florida ^73^ | The University of Florida health precision medicine program led the clinical implementation of alerts for six gene-drug pairs. Lessons learned include prescriber education methods, clear concise guidance on genotype-based actions, and real time clinical results are key. | Clinicians in USA | Preemptive PGx testing alerts clinicians at point of care |
| Other CDS for precision medicine | | | |
| Way, H.; 2021; Australia ^74^ | Implementation review for IntellxxDNA, a tool used to evaluate genetic data and return a genomically targeted report based on specific SNP's, improved care for patients | Clinicians in Australia | Collect information about specific SNP's and return in a report to the clinician (genomically targeted intervention strategies including nutrients, supplements, and lifestyle modifications) |
| Baye, J.F.; 2020; South Dakota ^75^ | Malignant hyperthermia susceptibility: Utilization of genetic results in an electronic medical record to increase safety | Clinicians in USA | Alert system for malignant hyperthermia susceptibility |
| Edelman, E.A.; 2013; Maryland ^76^ | Evaluation of a novel electronic genetic screening and CDS tool in prenatal clinical settings | Clinicians and patients in USA | Computerized intervention for identification and management of prenatal genetic risks using FHx |
| Reumkens, 2020; Netherlands ^77^ | Clinical surveys of decision aid to support couples during reproductive decision-making found barriers and facilitators to implementation | Clinicians and patients in Netherlands | Online decision aid used to supplement counseling in a targeting group to make reproductive decisions |
| Marwaha, 2021REF; Face2Gene ^78^ | System description and implementation review for Face2Gene, which was designed to use facial phenotyping to suggest a potential diagnosis | Clinicians in Canada | Phenotype driven prediction of genetic diagnosis based on facial features |

CDS, Clinical decision support; FHx & FHH, family health history; PCP, primary care physician; GPM, genetically guided precision medicine; EHR, electronic health record, FHIR, Fast Healthcare Interoperability Resources; HL7, Health Level Seven International; SNP, Single Nucleotide Polymorphis; BPA, Best Practice AlertAdvisory; RE-AIM: Reach, effectiveness, adoption, implementation, and maintenance framework;
